# Metric selection and promotional language in health artificial intelligence

**DOI:** 10.1101/2021.09.27.21264169

**Authors:** S. Scott Graham, Trisha Ghotra

## Abstract

**Background:** Recent advances in Artificial intelligence (AI) have the potential to substantially improve healthcare across clinical areas. However, there are concerns health AI research may overstate the utility of newly developed systems and that certain metrics for measuring AI system performance may lead to an overly optimistic interpretation of research results. The current study aims to evaluate the relationship between researcher choice of AI performance metric and promotional language use in published abstracts.

**Methods and findings:** This cross-sectional study evaluated the relationship between promotional language and use of composite performance metrics (AUC or F1). A total of 1200 randomly sampled health AI abstracts drawn from PubMed were evaluated for metric selection and promotional language rates. Promotional language evaluation was accomplished through the development of a customized machine learning system that identifies promotional claims in abstracts describing the results of health AI system development. The language classification system was trained with an annotated dataset of 922 sentences. Collected sentences were annotated by two raters for evidence of promotional language. The annotators achieved 94.5% agreement (κ = 0.825). Several candidate models were evaluated and, the bagged classification and regression tree (CART) achieved the highest performance at Precision = 0.92 and Recall = 0.89. The final model was used to classify individual sentences in a sample of 1200 abstracts, and a quasi-Poisson framework was used to assess the relationship between metric selection and promotional language rates. The results indicate that use of AUC predicts a 12% increase (95% CI: 5-19%, p = 0.00104) in abstract promotional language rates and that use of F1 predicts a 16% increase (95% CI: 4% to 30%, p = 0. 00996).

**Conclusions:** Clinical trials evaluating spin, hype, or overstatement have found that the observed magnitude of increase is sufficient to induce misinterpretation of findings in researchers and clinicians. These results suggest that efforts to address hype in health AI need to attend to both underlying research methods and language choice.

## Introduction

Popular and scientific accounts describing the potential of Artificial Intelligence (AI) for health and medicine promise fundamental transformations in the nature and quality of care.^1,2^ Accounts of the near future of health AI promise full life-span benefits from reproductive planning through end-of-life care.^1-2^ as well as transformation in related areas like health policy.^3^ Recently developed and currently available health AI technologies have shown great potential for cancer diagnosis,^3^ intensive care unit admission prediction,^4^ health policy,^5^ and even mitigating systemic biases in medicine.^6^ These kinds of promising results are fueling unprecedented investments in the sector, with over $14 billion in US venture capital in 2020.^7^ The enthusiasm for health AI is often warranted. However, many in medicine and bioethics are concerned that if practitioners or hospital systems put too much trust in health AI’s promotional claims, it may lead to significant patient harm.^7-13^ Overly enthusiastic adoption of AI may lead to misdiagnoses, medical errors, and inequitable delivery of care.

To help address these issues, researchers in medicine and bioethics are advancing new standards for health AI research and reporting.^14-16^ Nevertheless, the ability to effectively identify extravagant and promotional claims will remain an important part of vetting new technological innovations. Appropriate validity methods and related performance metrics are often seen as the cornerstone of these initiatives. The ability to precisely determine the accuracy of a given AI system offers an important tool for separating the hype from reality, in terms of both AI performance and potential utility. Specificity and sensitivity, for example, have been primary metrics in diagnostic medicine for nearly 75 years.^17^ These metrics are relatively intuitive ratios confusion matrix values (true positives, true negatives, false positives, false negatives). Specificity is defined by the number of true negatives over the sum of true negatives and false positives; whereas, sensitivity represents the number of true positives over the sum of true positives and false negatives. In short, these metrics offer clinicians critically important information about how likely a given test is to correctly diagnose a patient and how likely that same test is to correctly clear a patient. Other common metrics such as accuracy, precision, and recall are derived similarly based on a simple confusion matrix. However, health AI researchers increasingly use composite metrics that mathematically aggregate these ratios. Area Under the Receiver Operating Characteristic Curve (AUROC or AUC) and F1 are among the most popular. AUC is determined by plotting a ROC curve defined by sensitivity and 1-specificity and then calculating the two-dimensional area under that curve. F1 is the harmonic mean of precision and recall. While composite metrics can be helpful when comparing the performance of different candidate models built under the same framework, many in clinical medicine have expressed concerns that these metrics are often misunderstood and do not offer healthcare providers critically important information about diagnostic performance.^18-20^ The underlying question of this study, therefore, is to evaluate if use of these composite metrics might associate with the kinds of increases in promotional language that lead to overconfidence in new AI systems.

## Methods

To evaluate this question, we assessed the relationship between metric selection and promotional language usage rates in a random sample of 1200 abstracts collected from PubMed. Promotional language was identified using a custom machine learning classifier trained on human-annotated sentences from previously collected abstracts reporting the results of newly developed health AI systems. In what follows, we describe our search strategy and sampling technique. Subsequently, we describe the development and validation of our promotional language classifier.

### Search strategy and data collection

Our goal in this study was to curate a dataset that would allow us to assess any potential relationship between use of composite metrics and rates of promotional language related to health AI. Specifically, our aim was to collect research on health-related machine learning systems that included relevant benchmarking data in the PubMed-indexed abstract. In order to do so, we began by implementing a search strategy that identified PubMed-indexed articles containing (1) “machine learning” within the Medical Subject Heading (MeSH) ontology, and (2) one of the following terms in either the published article title or abstract: Accuracy, AUC, AUROC, F1, F-1, Negative Predictive Value, NPV, Positive Predictive Value, PPV, Precision, Recall, Sensitivity, Specificity, TNR, TPR, True Negative Rate, or True Positive Rate. This search yielded a total of 15,481 papers. Collected papers were subsequently screened for English language, presence of an abstract, and discussion of specific metrics within the abstract. Since the available PubMed search protocol bundles the title and abstract fields, this secondary screening focused on abstracts alone was necessary to locate papers that fit the study inclusion criteria. 7,421 records remained after screening, and a random sample of 1200 was extracted for subsequent analysis. This sample size was identified prospectively in order to assure 95% power.^21^ See Fig. 1 for additional details on the search strategy and selection of articles.

**Fig. 1.**
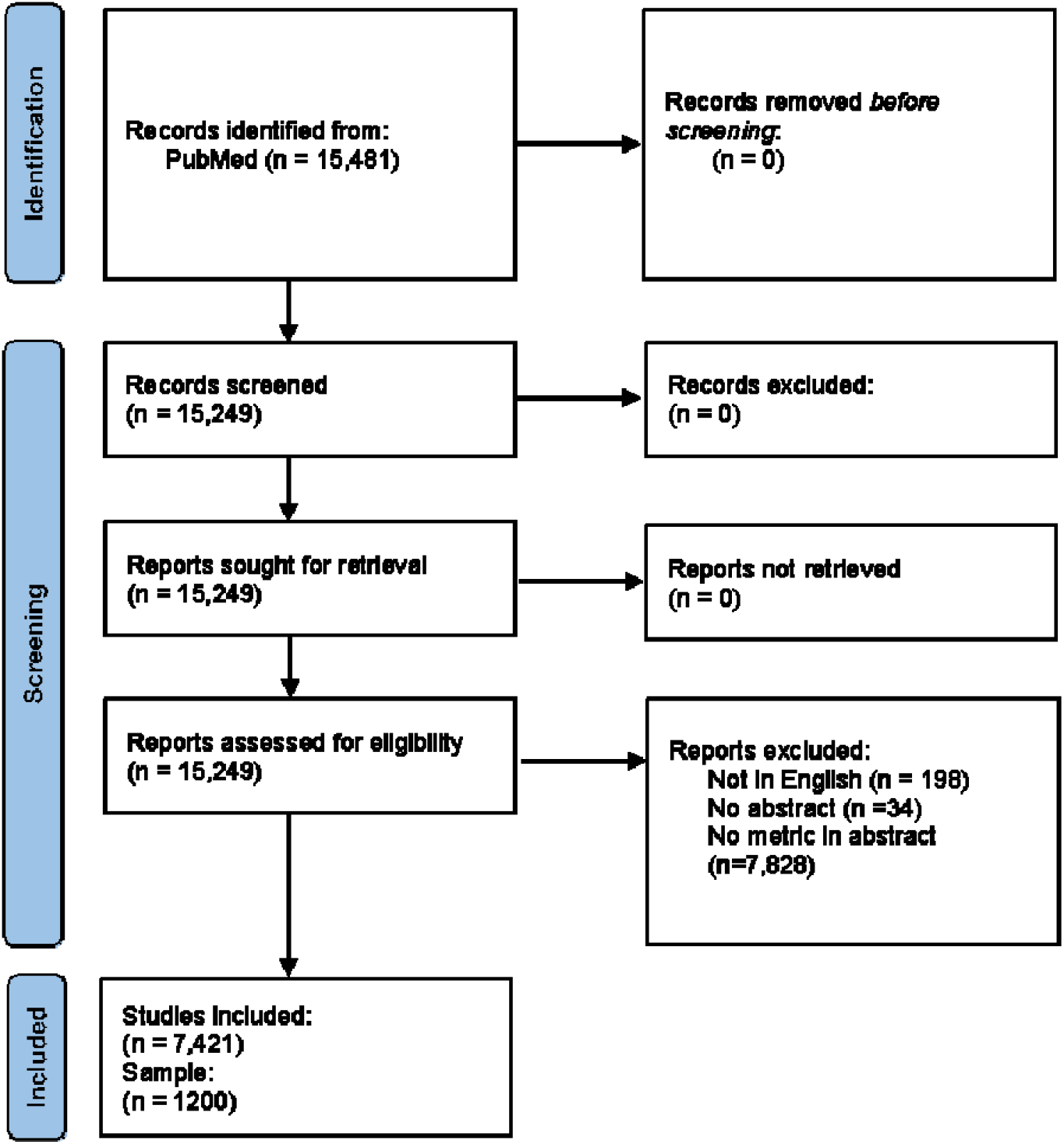
Identification, Screening, and Inclusion of Studies. This flowchart details the identification, screening, and inclusion of studies for this paper. The chart is an adaptation Adapted from the PRISMA 2020 Guidelines.^22^

For each abstract in the sample, we collected data on which and how many performance metrics were being used to evaluate tested systems. Since AUC and F1 are composite metrics, they are often used alongside their constituent metrics. Each new metric presented in an abstract, therefore, affords writers the opportunity to use more promotional language. Subsequently, it was important to know if the number of metrics alone was sufficient to predict changes in the relative frequency of promotional language use. Metric identification was based on the calculation or framework rather than simply the name of the metric. So, for example, instances of true positive rate (TPR), sensitivity, and recall were all identified as TPR since these metrics use identical calculations. Table 1 details the regular expressions used to identify and classify metrics in each abstract.

**Table 1.**
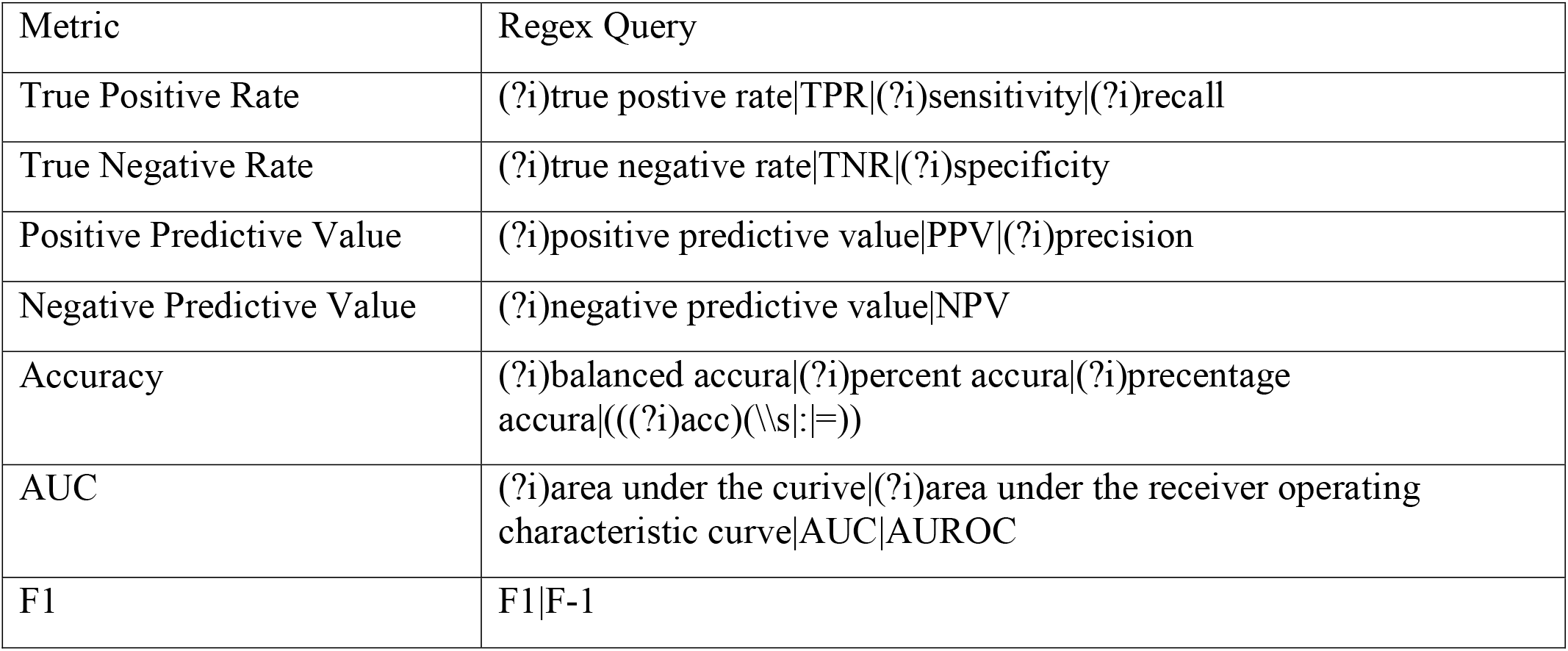
Metric identification and classification regular expressions.

### Annotation and Model Training

For the promotional language analysis, we built a natural language processing machine learning model capable of identifying promotional claims in the collected abstracts. The model was trained on human identification of promotional and non-promotional statements in research abstracts for 82 articles collected as part of a pre-existing meta-study on diagnostic AI performance.^23^ The meta-study inclusion criteria selected for articles that (1) evaluated a diagnostic classification task for a specific disease, (2) used deep learning models, and (3) compared system performance to healthcare professionals. The study authors qualitatively evaluated 82 publications and included 25 in the statistical meta-analysis. Collected articles include evaluations of AI systems designed to support diagnostic imaging in oncology, dermatology, ophthalmology, cardiology, and other assorted subspecialties. To curate the training set, we searched PubMed for each article in the original meta-study and collected available abstracts. Seventy-six of the abstracts are publicly available via PubMed. The remaining six abstracts were collected directly from publisher websites.

Collected abstracts were tokenized by sentence creating a dataset of 922 sentences for annotation. Each of these sentences was annotated by two annotators for evidence of promotional claims about developed AI system(s). The promotional language annotation was assigned when one of the following were present: (1) favorable comparisons to human annotators or previously developed health AI systems, (2) positive superlative qualifying adjectives describing the performance or efficiency of the system, (3) claims to generalizability or clinical applicability, or (4) assertions that system performance meets the standards for Food and Drug Administration medical device clearance or approval. Many sentences classified as promotional met multiple requirements for annotation. For example, it was common for favorable comparisons to include superlative qualifiers. A given sentence was only annotated as “promotional language” if it described a system under evaluation in the article. Promotional language about AI, in general, was not assigned to the “promotional” category. Additionally, claims about system performance that might be considered objectively good (e.g., AUC=.9997) were not classified as “promotional” unless the sentence also included favorable comparisons, positive superlative qualifiers, claims to generalizability/applicability, or claims to meeting regulatory standards. Further details about these features and illustrative examples are available in Table 2.

**Table 2.**
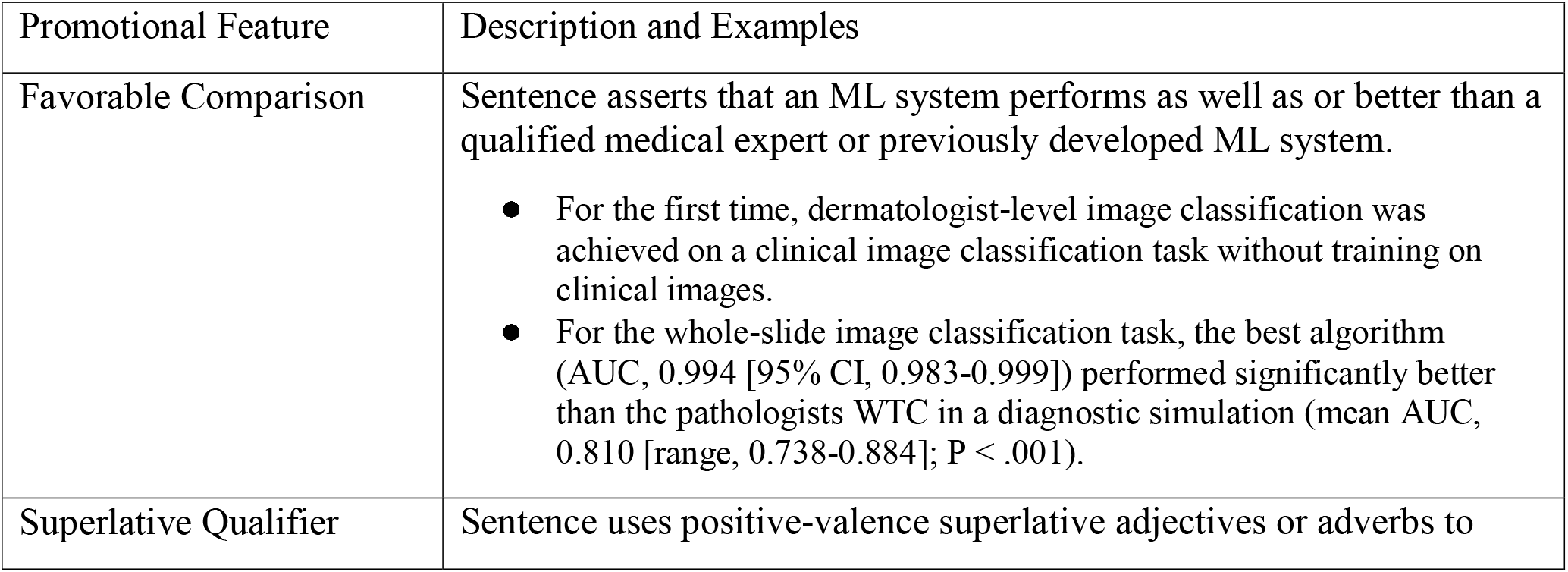
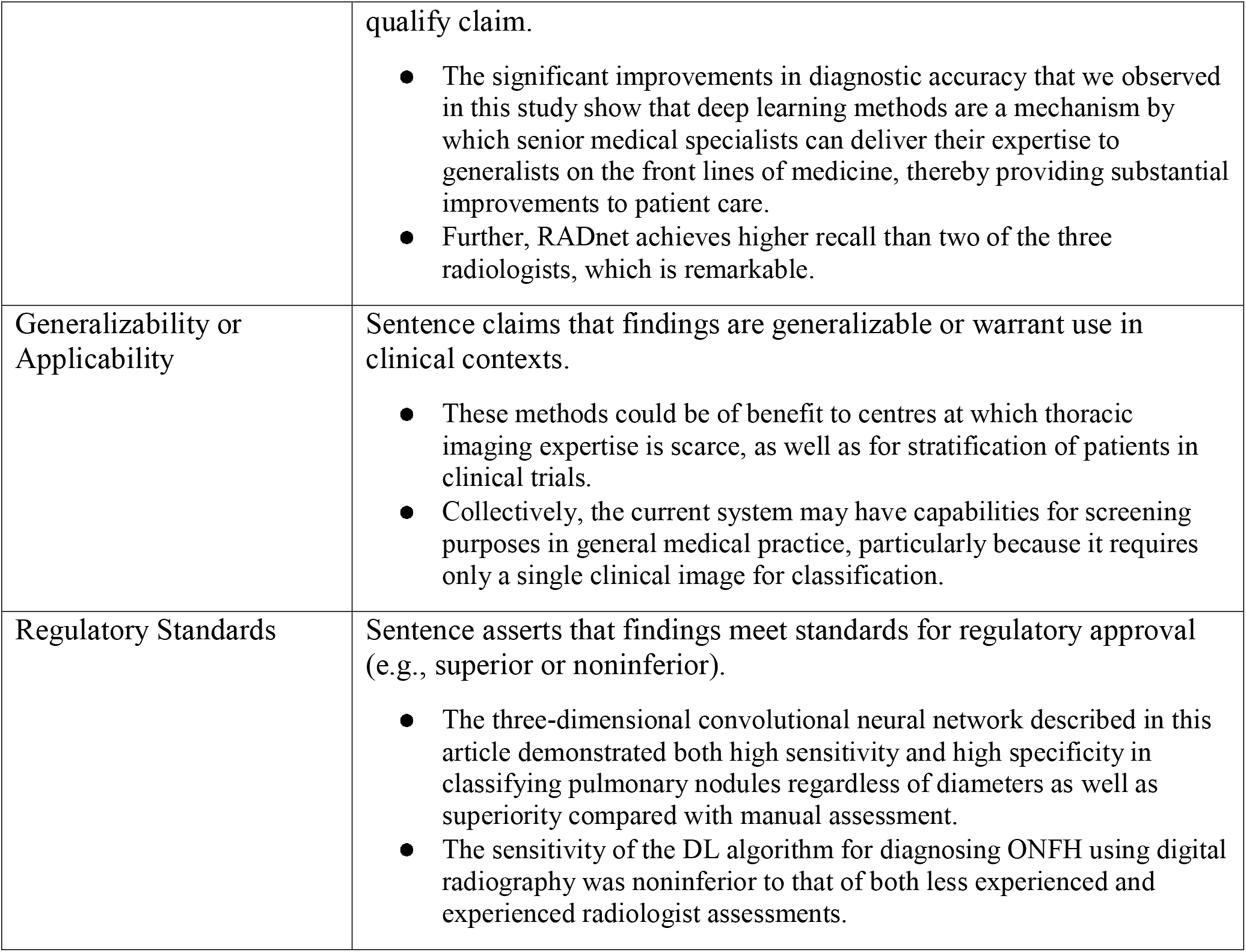
Common features of promotional language and illustrative examples.

Initial annotation was completed on a subsample of 288 sentences. Annotations were applied independently by each annotator and inter-rater reliability was assessed using Cohen’s kappa. Initial inter-rater agreement was 94.1% (κ = 0.812). The two annotators used these results to conduct an additional norming exercise where they discussed points of disagreement. They then re-annotated the original 288 sentences independently, along with the remaining 634 sentences to create the final dataset. Final interrater reliability was almost perfect (κ = 0.825 with 94.5% agreement) according to previously published guidelines for qualitative interpretation of Cohen’s κ.^24^ The few remaining annotation disagreements were resolved by a third annotator prior to model training.

We extracted relevant features from the annotated sentences and assessed several competing approaches to training in order to develop the final model. All feature extraction and training was implemented in R version 4.0.2, although some feature extraction techniques made use of python virtual environments via the reticulate library.^25^ All feature extraction and modeling was performed on a desktop workstation (Dell G5, core i3-9300, 16 GB RAM). Part-of-speech (POS) average location (aveloc) was used for primary feature extraction.^26^ POS aveloc uses spaCyr to classify each word according to POS type and identifies the average location within each sentence.^27^ The method was designed to provide a purely syntactic approach to feature engineering in a computational social science context.^26^ For POS aveloc to work well, it generally requires cases where (1) the unit of analysis is a sentence, (2) the collected sentences are fairly homogeneous in terms of content, and (3) the label of interest is a discursive or linguistic strategy/technique. POS aveloc feature engineering occurs in two steps: (1) The text of interest is parsed to identify parts-of-speech, and (2) the average location (within each sentence) of each part of speech is identified. POS aveloc feature engineering was augmented with a sentence order variable that resulted in moderate increases in accuracy. Candidate models included k-nearest neighbor (KNN), Bagged classification and regression trees (CART), naïve bayes, and neural network (NNet). All training was implemented in caret using a random 80/20 train/test split and 10-fold cross-validation.^28^ The bagged CART models had the highest accuracy across metrics. Performance, recall, and AUC measures are available in Table 3, and the ROC curves for the final models are available in Figure 2.

**Table 3.** Precision, recall, and AUC for candidate models.

| **Model** | **Precision** | **Recall** | **AUC** |
| --- | --- | --- | --- |
| Bagged CART | 0.92 | 0.89 | 0.8947 |
| NNet | 0.86 | 0.85 | 0.7685 |
| KNN | 0.84 | 0.96 | 0.7501 |
| Naive Bayes | 0.8 | 0.944 | 0.7614 |

**Fig. 2.**
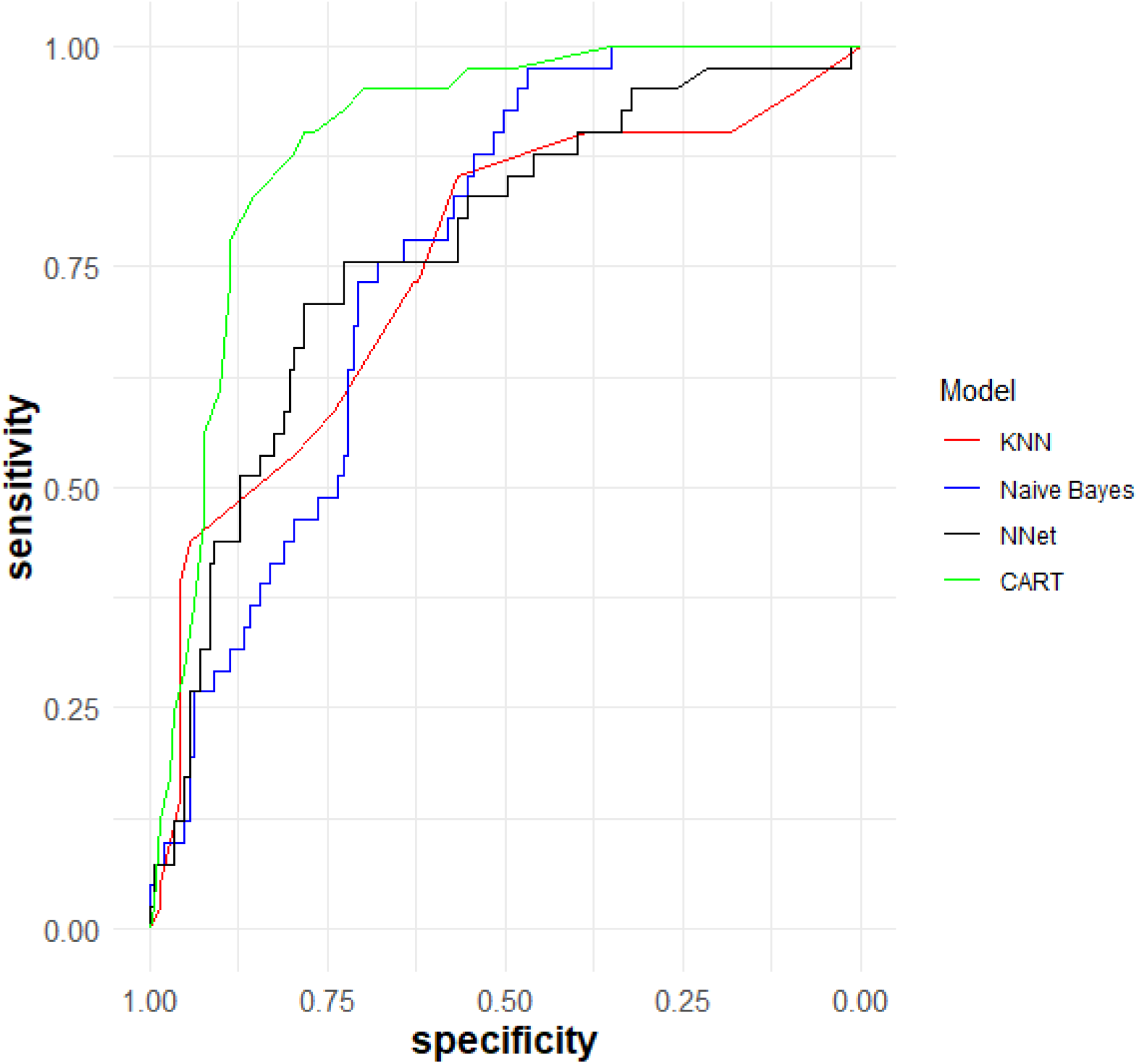
ROC curves for candidate models. Figure shows the differential performance of KNN (red), Naive Bayes (blue), Neural Net (black), and bagged CART (Green) classification models. ROC curves were generated with the pROC R library.^29^ The bagged CART model was the most performant with a total area under the curve of 0.8947.

## Results

The 1200 studies included in this study focused on the development or validation of health AI systems in a wide variety of clinical areas. The studies were all published between 2010 and 2021 in 421 distinct journals. The most commonly represented journals included *Scientific Reports* (58 articles), *PLOS One* (56 articles), *Computers in Biology and Medicine* (36 articles), *Annual International Conference of the IEEE Engineering in Medicine and Biology Society* (31 articles), *Sensors* (27 articles), *Computer Methods and Programs in Biomedicine* (23 articles), *European Radiology* (23 articles), *BMC Bioinformatics* (18 articles), *BMC Medical Informatics and Decision Making* (15 articles), and the *International Journal of Medical Informatics* (13 articles).

Each evaluated abstract reported between 1 and 5 metrics, with an average of 1.66. The number of sentences identified as containing promotional language ranged from 0 to 12, with an average of 2.67. Journal conventions for abstract length vary widely with unstructured abstracts often being as few as five sentences and structured clinical abstracts sometimes being as many as 20. Therefore, we used the promotional language flagged sentences to determine the percentage of promotional sentences in each abstract. The proportion of promotional claims in each abstract ranged from 0 to 60% with an average of 22.85%, slightly higher than the 20% norm identified in linguistic studies of biomedical abstracts.^30^ Table 3 provides additional details regarding distribution of promotional statements and number of metrics per abstract.

**Table 3:** Distribution of promotional statements and number of metrics.

|  | Low | Mean (SD) | High |
| --- | --- | --- | --- |
| Promotional Statements (N) | 0 | 2.67 (1.89) | 12 |
| Promotional Statements (%) | 0 | 22.85 (13.1) | 60.0 |
| Number of Metrics | 1 | 1.66 (0.79) | 5 |

To evaluate the relationship between use of AUC, use of F1, total number of metrics, and the proportion of promotional sentences, we used a quasi-Poisson framework. While both use of AUC and F1 proved to be significant predictors of promotional language use (p = 0.00149 and p = 0.01564), the number of metrics used was not a significant predictor (p = 0.53216), So we removed the number of metrics variable from the model. The final model predicts a 12% increase (95% CI: 5-19%, p = 0.00104) in the promotional language rates for abstracts that report AUC and a 16% increase (95% CI: 4% to 30%, p = 0. 00996) for abstracts that use F1. See fig. 3 for additional details.

**Fig. 3.**
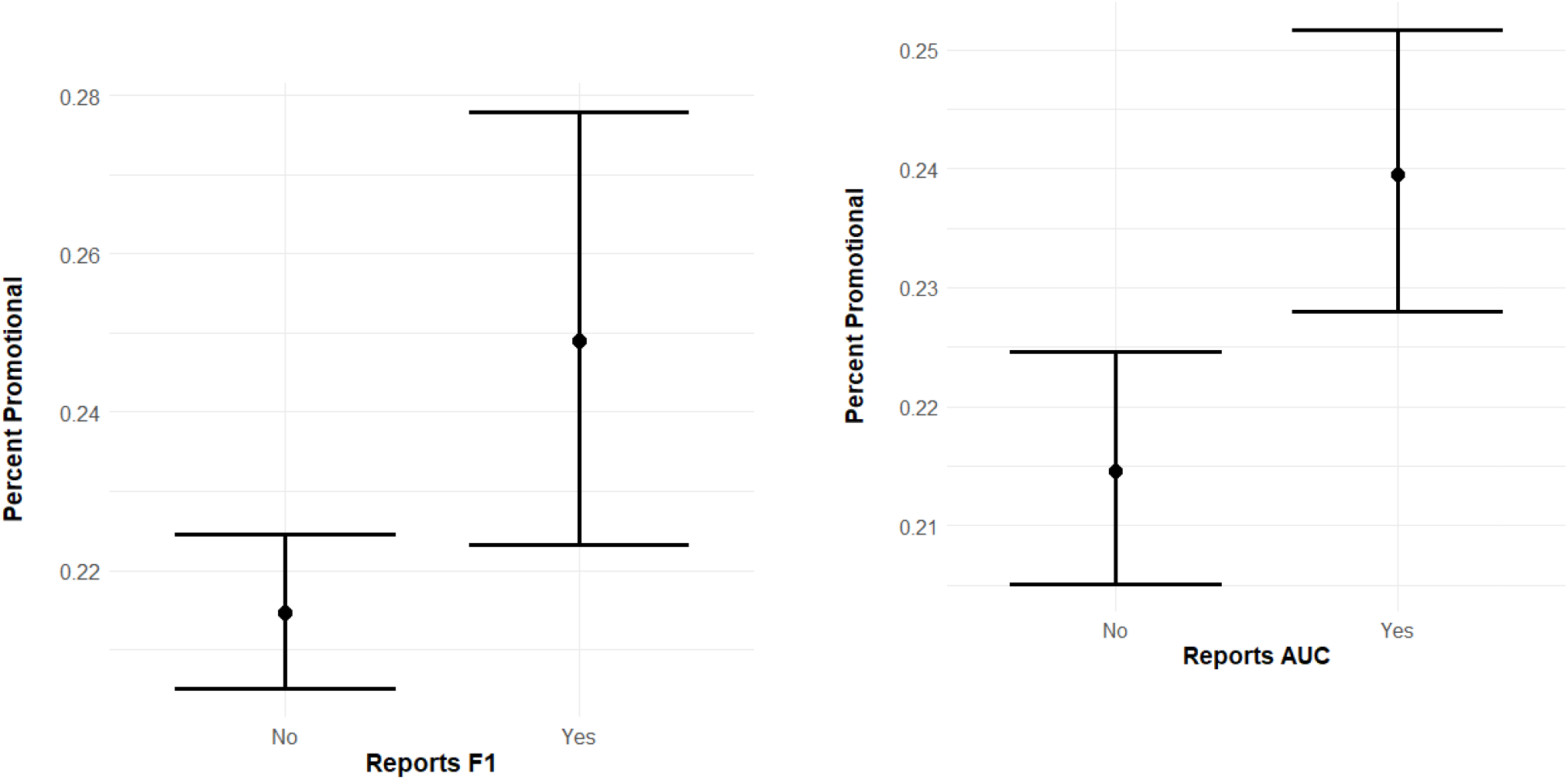
Promotional language rates by F1 (left) and AUC (right) usage. Figure details the incident rate ratios and 95% confidence intervals for the proportion of promotional language in each abstract by composite metric usage.

## Discussion

While the overall observed magnitude of the increase is not large, it is consistent with promotional language differentials seen in randomized clinical trials of researcher and clinician responses to spin or overstatement. In one study, a single sentence of overstatement was enough to prompt improper assessment of research results in 44.4% of cases.^31^ Importantly, the effect of an overstated sentence was not the same across reader demographics. While clinicians who had led research projects were less likely to accept a face-value reading of an overstated claim, time since graduation for practicing clinicians predicted a greater likelihood of accepting overstated claims. Modest increases in promotional language have also been shown to lead researchers and clinicians to be more likely to evaluate new treatments as potentially beneficial to patients, even though the presence of spin led researchers to rate studies as less methodologically rigorous.^32^

Advances in machine learning and AI have the potential to substantially improve biomedical research and clinical practice. However, the adoption of new AI innovations that do not live up to the promises made in research reports can lead to both adverse events for patients and overall distrust in the potential benefits of clinical AI. Excessive promotional language, hype, or spin helps to create the conditions for these adverse outcomes. Efforts to study and address promotional language use in scientific and biomedical research tend to focus on hype in popular media.^33-36^ Within this framework, much of the research on hype or overstatement evaluates mismatches between the underlying research and the presentation of findings in press releases and news articles. Similar research also evaluates inconsistencies between research results and the presentation of findings in abstracts or published articles.^20-21^ These common methodologies for addressing hype assume a readily identifiable division between the underlying research and the linguistic presentation of results. The results presented in this article indicate that some methods themselves may lead to measurable increases in promotional language. Subsequently, these findings suggest that efforts to address hype in health AI need to attend to both underlying research methods and language choice in the presentation of findings. Given the established threats to clinical utility and the results of this study that indicate use of composite performance metrics can increase promotional language rates, health AI researchers and editorial boards may wish to reconsider ideal reporting practices in these areas. While composite metrics are quite useful when it comes to comparing the performance of candidate models within a study, authors and editors should be on guard against the attendant risks of increased promotional language that comes with the use of these metrics.

## Data Availability

Data are available upon request.

## Acknowledgments

S.S.G. and T.G. thank Vivian Tran for her assistance with promotional language annotations.

